# Tissue-specific microRNA signatures in pediatric inflammatory bowel disease with juvenile idiopathic arthritis: differential expression and cross-platform evaluation

**DOI:** 10.1101/2025.11.06.25339736

**Authors:** Maria Dorn-Rasmussen, Jaslin P James, Mikkel Malham, Thea A Hvidtfeldt, Lone Schejbel, Boye S Nielsen, Johan Burisch, Helene A S Ingels, Estrid V S Høgdall, Lene B Riis, Vibeke Wewer

**Author notes:** CORRESPONDENCE Maria Dorn-Rasmussen, Department of Paediatric and Adolescent Medicine Copenhagen University Hospital, Amager and Hvidovre Kettegaard Allé 30, DK-2650 Hvidovre, Denmark. **SPECIFIC AUTHOR CONTRIBUTIONS:** VW and LBR conceptualized the study. MDR, JPJ, MM, BSN, JB, HASI, EVSH, LS, LBR, and VW collectively planned and designed the research. MDR was primarily responsible for data acquisition and conducted the RNA sample extraction, microRNA microarray hybridization, washing, staining, and scanning procedures in the discovery study. JPJ executed the statistical analyses of microarray data. TAH conducted the RT-qPCR validation. MDR prepared the initial draft of the manuscript. All authors had complete access to the data throughout the study and participated in critically revising the manuscript to ensure it met rigorous intellectual standards. Each author has approved the final manuscript draft. The decision to submit the manuscript for publication was made jointly by LBR and VW. The corresponding author confirms that all individuals listed as authors meet the authorship criteria, with no omissions of individuals meeting these criteria.

## Abstract

**Background & Aim:** Development of extraintestinal manifestations in pediatric inflammatory bowel disease (pIBD) is associated with a more severe disease course; juvenile idiopathic arthritis (JIA) is the most frequent extraintestinal manifestation in pIBD. We investigated whether intestinal mucosal microRNAs measured at pIBD diagnosis could predict subsequent JIA (pIBD-JIA) before symptoms emerge.

**Methods:** We analyzed formalin-fixed, paraffin-embedded diagnostic intestinal biopsies from 101 pediatric patients under 18 years of age: a discovery cohort (n=62; 27 pIBD, 25 pIBD-JIA, 10 non-IBD controls) and a validation cohort (n=39; 29 pIBD, 10 non-IBD; pIBD-JIA cases reused from discovery cohort). Discovery profiling used Affymetrix GeneChip™ miRNA 4.0. Significant probe sets were cross-referenced in MirGeneDB 2.1 and literature-curated to nominate candidates for RT-qPCR validation.

**Results:** We identified 532 differentially expressed probe sets across groups (adjusted p<0.05); 123 microRNAs showed absolute log₂ fold change >2. Thirteen microRNAs exhibited the clearest separation between pIBD and pIBD-JIA; 11 achieved AUC>0.70 in discovery. Four candidates (miR-663a, miR-3197, miR-4463, miR-92a-3p) were advanced to RT-qPCR validation. Two assays (miR-3197, miR-4463) failed technical criteria; differences in miR-663a and miR-92a-3p were reproduced within a discovery subset but did not replicate in the pIBD/non-IBD validation cohort.

**Conclusions:** Diagnostic mucosal microRNA profiles distinguished pIBD patients who later developed JIA in discovery, but selected microRNA candidates were not general across cohorts. These findings underscore both the potential of tissue microRNAs as prognostic markers and the need for larger, fully independent, standardized studies before clinical translation.

## 1. INTRODUCTION

Inflammatory bowel diseases (IBD) are chronic inflammatory conditions of the gastrointestinal tract, including Crohn’s disease (CD), ulcerative colitis (UC), and IBD-unclassified. The global incidence of IBD continues to rise, and Denmark ranks among the countries most affected in terms of prevalence.^1,2^ IBD is frequently diagnosed in childhood or early adolescence. Pediatric patients with IBD (pIBD) often experience a more severe disease course compared to adults, characterized by a more extensive disease, frequent disease flares, hospitalizations, growth retardation, and an increased need for early advanced therapies and surgery.^3,4^ Up to one third of patients with pIBD develop extraintestinal manifestations (EIMs), which is associated with a more severe disease course.^5–7^ An EIM in IBD is defined as *an inflammatory pathology in a patient with IBD that is located outside the gut and for which the pathogenesis is either dependent on extension/translocation of immune responses from the intestine, or is an independent inflammatory event perpetuated by IBD or that shares a common environmental or genetic predisposition with IBD*.^8^ Most EIMS in IBD affects the joints, skin, eyes, or the hepatobiliary system. In pIBD, juvenile idiopathic arthritis (JIA) is the most common comorbidity affecting approximately 16–33%.^9,10^ JIA is anautoimmune disease which causes chronic joint inflammation. JIA occurs, by definition, before the age of 16 and poses significant risks for patients with pIBD, including uveitis and long-term complications such as reduced sight, joint destruction, reduced physical activity, and psychological challenges.^11^ Despite advancements in diagnostics and treatment, a reliable biomarker for predicting JIA in pIBD remains an unmet clinical need.^12^

The underlying pathogenesis of pIBD and JIA remains incompletely understood, despite extensive research.^13^ Genetic susceptibility, epigenetic modifications, environmental exposures, and alterations in the gut microbiota are recognized as key contributors to disease progression in pIBD.^13–16^ In recent years, particular attention has been directed towards epigenetic regulatory mechanisms, including the role of microRNAs (miRNAs).^17^

MiRNAs are small, endogenous, non-coding RNA molecules, typically around 22 nucleotides in length, that regulate gene expression at the post-transcriptional level.^18^ They are involved in a wide range of cellular processes, including the proliferation, maturation, and differentiation of immune cells such as T and B lymphocytes, thereby playing a central role in the modulation of immune responses.^19^ Dysregulated miRNA expression has been associated with several immune-mediated inflammatory diseases, including IBD and JIA.^20^ While extensive research has been conducted on miRNAs in adult IBD, their role in pIBD remains poorly characterized.^21–23^

During the past decade, miRNAs have recently been recognized as potential biomarkers in both IBD and JIA.^24,25^ Their tissue-specific expression and stability in biological samples further enhance their suitability for biomarker applications, particularly in intestinal biopsies.^26^ Technological advances, including microarrays and reverse transcription quantitative polymerase chain reaction (RT-qPCR), have enabled more precise quantification of miRNA expression, thereby providing valuable insights into disease mechanisms. Identifying disease-specific miRNA signatures could enhance our understanding of pIBD and pIBD who later develop JIA. This may facilitate the development of biomarkers with diagnostic and prognostic utility.^26^

In this study, we aimed to identify miRNA biomarkers in patients with pIBD that predict subsequent development of JIA, using multiple analytic assays and platforms.

## 2. MATERIALS AND METHODS

### 2.1. Study design

This cross-sectional observational study investigated differences in miRNA expression levels between patients with pIBD, patients with both pIBD and JIA (pIBD-JIA), and non-IBD controls. The study consisted of a discovery study, where differentially expressed miRNAs were identified using microarray analysis, followed by a validation study, where selected miRNA candidates for distinguishing patients with pIBD and patients with pIBD-JIA, were assessed by RT-qPCR. Formalin-fixed, paraffin-embedded (FFPE) tissue samples were retrospectively selected from the index endoscopy performed as part of the diagnostic workup for pIBD. At the time of endoscopy, no patients had yet developed JIA.

Data were accessed for research purposes between January 2022 and November 2025. Only Dr. Maria Dorn-Rasmussen, had access to potentially identifiable participant information during or after data collection.

### 2.2. Study population

#### 2.2.1. Discovery study

This cohort included patients less than 18 years of age diagnosed with pIBD between 2010 and 2024 and who were treatment naïve. Among these, a subset of patients less than 16 years of age subsequently developed JIA (pIBD-JIA). Patients with systemic-onset JIA were excluded, as multiple studies indicate that systemic-onset JIA is biologically distinct and exhibits an inflammatory profile that differs from other JIA subtypes.^27,28^ A non-IBD control group less than 18 years of age undergoing clinically indicated endoscopy due to suspected pIBD was also included. Non-IBD controls all exhibited normal endoscopic and histopathological findings. All intestinal biopsies were collected during the diagnostic endoscopy for pIBD at Copenhagen University Hospital, Hvidovre. Accordingly, biopsies from pIBD patients who subsequently developed JIA were obtained before the onset and diagnosis of JIA. All patients resided within the catchment areas of the Copenhagen University Hospitals.

#### 2.2.2. Validation study

The validation cohort consisted of group of treatment naïve patients less than 18 years of age with pIBD and a non-IBD control group identified at the same hospital center during 2002–2020. All intestinal biopsies were collected during the diagnostic endoscopy at Copenhagen University Hospital, Hvidovre. For the IBDJIA subgroup, validation analyses were performed on the same patients as in the discovery cohort; therefore, the IBDJIA validation represents an internal (non-independent) validation. Patients in the validation cohort were selected to resemble those in the discovery cohort with respect to diagnosis, sex, and age, however, complete matching was not feasible due to the limited number of available biopsies

### 2.3. Samples and data collection

#### 2.3.1. Samples

FFPE mucosal biopsies from the terminal ileum, caecum, ascending colon, descending colon, sigmoid colon, and rectum were routinely collected from all patients during their index endoscopy, as part of the standard diagnostic workup for pIBD. Histopathological evaluation of all tissue sections stained with hematoxylin and eosin (H&E) was conducted by two board certified pathologists (Dr. Lene B Riis & Dr. Eva Gravesen) from the Department of Pathology, Copenhagen University Hospital, Herlev and Gentofte. Based on this assessment, the biopsy exhibiting the most pronounced inflammatory changes was carefully selected from each patient to ensure consistency in sampling and to capture the area of maximal inflammation.

#### 2.3.2. Data collection for multivariate analysis

Demographic characteristics, including sex and age, were collected for all patients. Patients were divided into three groups according to age at diagnosis: A1a, 0–<10 years; A1b, 10–<17 years; and A2, 17–40 years (with all patients aged 17 years included) in accordance with the Paris classification.^29^ Clinical and endoscopic disease activity at diagnosis was assessed using the abbreviated Pediatric Crohn’s Disease Activity Index (abbrPCDAI)^30^ and Simple Endoscopic Score for Crohn’s Disease (SES-CD)^31^ for patients with pediatric Crohn’s disease (pCD) and the Pediatric Ulcerative Colitis Activity Index (PUCAI)^32^ and Mayo Endoscopic Subscore (MES)^33^ for patients with pediatric ulcerative colitis (pUC). Biochemical assessments at diagnosis were also collected: C-reactive protein (CRP), hemoglobin, albumin, fecal calprotectin. Histopathological disease activity graded using the Robarts Histopathology Index (RHI)^34^ was evaluated on H&E-stained slides from all selected biopsies from both pCD and pUC patients in the discovery cohort by a blinded pathologist (Dr. Lene B Riis), without knowledge of endoscopic or clinical activity scores.^35,36^ Sample location was divided in left- and right colon.

### 2.4. Definitions

pIBD diagnoses were established according to the revised Porto criteria, and all patients were classified using the Paris classification.^29,37^ JIA was diagnosed by pediatric rheumatologists based on the International League of Associations for Rheumatology (ILAR) classification system.^38^ Clinical remission in pIBD was defined as abbrPCDAI or PUCAI <10, mild disease activity as abbrPCDAI 10-27.5 or PUCAI 10–34, moderate disease activity as abbrPCDAI 30-37.5 or PUCAI 35–64, and severe disease activity as abbrPCDAI ≥40 or PUCAI 65–85.

Left-sided biopsies included those from the descending and sigmoid colon, while right-sided biopsies comprised those from the terminal ileum, caecum, ascending, and transverse colon. Histological remission was defined as RHI 0–3, mild disease activity as RHI 4–9, and moderate-to-severe disease as RHI ≥9.^39^

### 2.5. Laboratory methods

#### 2.5.1. RNA extraction

Total RNA was isolated from two 10 μm-thick sections of FFPE tissue using the MagMAX™ FFPE DNA/RNA Ultra Kit (Applied Biosystems™), adhering to the manufacturer’s protocol. Total RNA concentration was measured using the Qubit RNA HS Assay kit (Thermo Fisher scientific). All RNA samples were stored at −80°C until subsequent use.

#### 2.5.2. Discovery microarray profiling

MiRNA expression levels were assessed in all samples from the discovery study using the GeneChip™ microarray platform in combination with the FlashTag™ Bundle (Affymetrix, CA, USA). Labelling was performed with the FlashTag Biotin HSR kit, utilizing 130 ng of total RNA in 8 μl of nuclease-free water, in accordance with the manufacturer’s instructions. The labelled RNA was hybridized onto GeneChip™ miRNA 4.0 microarrays and incubated at 48°C for 16–18 hours.

Following hybridization, the microarray chips were washed and stained using the GeneChip Fluidics Station 450, and scanning was conducted with an Affymetrix GeneChip 7G scanner. Post-scan analysis was performed using Transcriptome Analysis Console (TAC) software, which includes initial data normalization, expression summarization, and differential expression analysis. All analyses were further reviewed and verified by an expert at ThermoFisher, ensuring the quality and reliability of the results obtained.

#### 2.5.3. RT-qPCR validation

Key miRNAs identified by microarray analysis, were selected for validation by RT-qPCR. Selection for validation was based on both statistical significance, fold change, and prior evidence from the literature, where these miRNAs had been implicated in IBD, JIA, or rheumatoid arthritis Universal reverse transcription (RT) of the miRNAs was conducted using the miRCURY LNA RT kit (Qiagen) as specified by the protocol. Briefly, RNA was diluted to 5 ng/µl and 2 µl was added to 8 µl reaction mix including 0.5 µl cDNA synthesis spike in control RNA (UniSp6). UniSp6 mixed with Total RNA control, Human (Thermo Fisher Scientific) was also used as the input in one cDNA reaction for later use in inter plate calibration (IPC). qPCR of the miRNAs of interest was performed using the miRCURY LNA SYBR Green kit (Qiagen) in combination with individual miRCURY LNA PCR assays (Qiagen). The individual assays include locked nucleic acid (LNA) miRNA-specific primers for the detection of the candidate miRNAs: miR-3197, miR-663a, miR-92a-3p and miR-4463, the reference miRNAs: miR-27a, miR-16-5p and mir-191-5p, the small nuclear RNA references: U6 and SNORD48 as well as UniSp6, used for detection of the spike in control and for IPC. Reactions were prepared as specified in the instructions using 60x dilutions of cDNA and were set up as 10 µl reactions in technical duplicates in 96 well plates (Roche). The qPCR was run with the following cycling conditions: 2 min heat inactivation (95°C) followed by 45 amplification cycles (95°C for 10 s and 56°C for 60 s) ending with a melt curve analysis. Cycle of threshold values (Ct values) were calculated using the absolute quantification/2^nd^ derivative maximum method of the LightCycler® 480 (Roche) software version 1.5.

### 2.6. Statistical Analysis

#### 2.6.1. Discovery study - Microarray data analysis

MiRNA expression data were pre-processed using the robust multichip average (RMA) algorithm, incorporating background correction, quantile normalization, and summarization.^40,41^ Probe sets with a log₂ expression intensity below 5 in more than 50% of samples were excluded to remove lowly expressed miRNAs and enhance the robustness of downstream analyses. Batch effects related to assay processing were corrected using the ComBat algorithm.^42^ Probe sets were filtered to include only *Homo sapiens*, and annotation was carried out using the pd.mirna.4.0 package.^43^ Differentially expressed miRNAs among IBD patients, those with both IBD and JIA, and non-IBD controls were identified using linear modelling implemented in the limma package. ^44^ Probe sets were ranked according to Benjamini-Hochberg adjusted p-values, which account for multiple testing, and the magnitude of fold changes in expression levels. The discriminatory ability of these miRNAs was evaluated by calculating the area under the receiver operating characteristic (ROC) curve (AUC), with significance defined as adjusted p-values below 0.05. To control for potential confounding factors, a multivariate analysis was performed, incorporating sex, age groups, clinical activity indices, histopathological disease activity, and sample location. Probe sets deemed significant were cross-referenced against the MirGeneDB 2.1 database to identify candidates for subsequent RT-qPCR validation.^45,46^ For each candidate miRNA, relevant biological pathways, molecular interactions, and curated literature links provided in MirGeneDB were reviewed to assess biological plausibility. In addition, complementary PubMed searches were conducted to identify studies reporting associations with inflammation, IBD, pIBD, arthritis, or JIA, thereby supporting the rationale for candidate selection. Microarray expression data have been deposited in ArrayExpress at the European Bioinformatics Institute (EMBL-EBI) under the accession number E-MTAB-15807. Analyses were performed in RStudio using R version 4.4.1.

#### 2.6.2. Validation study - RT-qPCR data analysis

Assays that did not exhibit good linearity (R^2^>0.98) or showed late Ct values (>38) were excluded for further analysis. Ct values of the UniSp6 spike in were used to ensure that RT was equally efficient across samples. Ct values of the UniSp6 IPC in the different runs were used to calculate a calibration factor used for Ct adjustment on every plate. Subsequently, averages of the technical duplicates were used in data analysis with the 2^-Δct^ method to calculate the relative expression of the candidate miRs. The average Ct value of the miR-27a, miR-16-5p, miR-191-5p, U6 and SNORD48 assays for each patient was used as the reference.^47^ To test whether significantly differential expression between patient groups (UC/UCJIA and CD/CDJIA) was present, a Welch t test was conducted. The data analysis was conducted in Microsoft Excel and R.

### 2.7. Ethical considerations

This study was approved by the Danish National Ethical Committee (reference number H-20065831) and the Danish Data Protection Agency (reference number P-2020-1065) as part of The Copenhagen IBD Inception Cohort Study, which commenced on 1 May 2021.^48^ All procedures were conducted in accordance with the ethical standards outlined in the Declaration of Helsinki and complied with both national and international guidelines.

## 3. RESULTS

### 3.1. Patient cohorts and characteristics

#### 3.1.1. Discovery cohort

The discovery cohort comprised 62 patients: 27 with pIBD (14 pediatric Crohn’s disease [pCD], 13 pediatric ulcerative colitis [pUC]), 25 with pIBD-JIA (10 pediatric Crohn’s disease and JIA [pCD-JIA], 15 pediatric ulcerative colitis and JIA [pUC-JIA]), and 10 non-IBD controls. **Figure 1** illustrates the study flowchart. Among patients with pIBD, the median age at diagnosis was 14 years (IQR: 12–16). In the IBDJIA group, the median age at IBD diagnosis was 12 years (IQR: 11–15) and 14 years (IQR: 12.5–16) at JIA diagnosis. Twenty-one (78%) and 19 (76%) of patients with pIBD and IBDJIA were classified as Paris A1b (10–<17 years), respectively. Detailed characteristics are shown in **Table 1** and stratified by disease subgroup in **Supplementary Table 1**.

**Figure 1.**
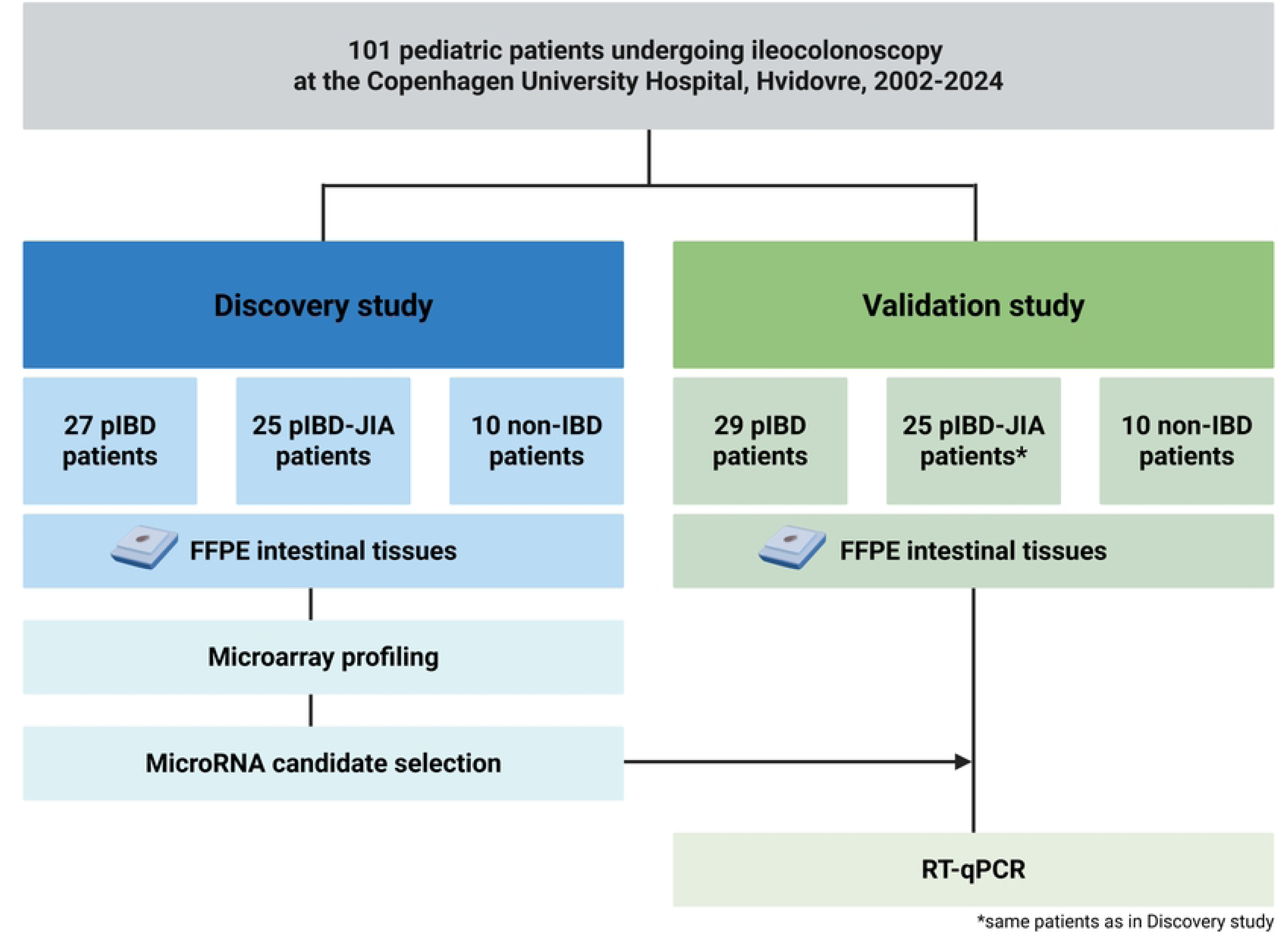
Study flowchart. pIBD, pediatric inflammatory bowel disease; pIBD-JIA, pediatric inflammatory bowel disease with later development of co-occurring juvenile idiopathic arthritis; non-IBD, non-inflammatory bowel disease; FFPE, formalin-fixed, paraffin-embedded; RT-qPCR, reverse transcription quantitative polymerase chain reaction.

**Table 1:**
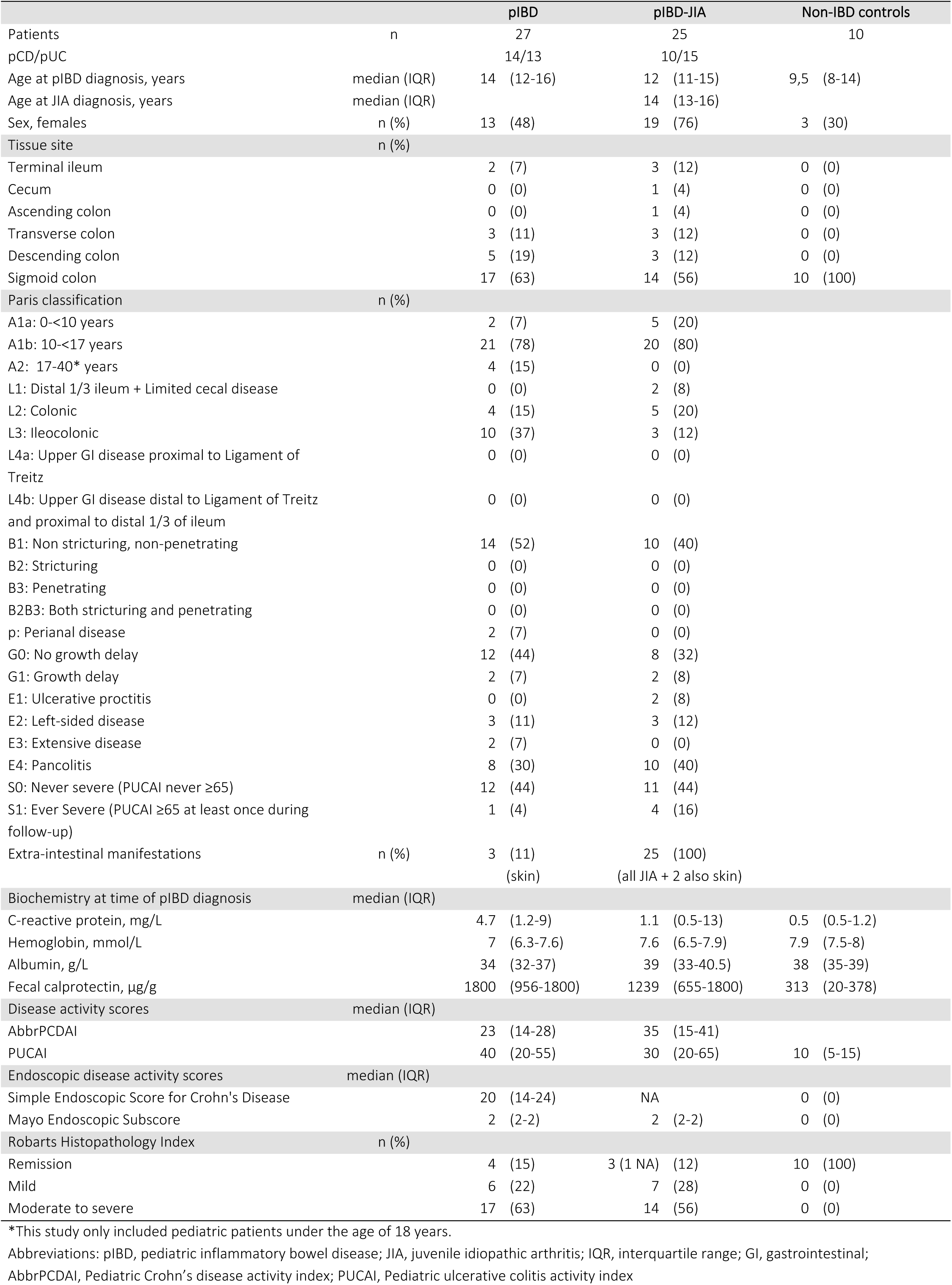
Patient characteristics of discovery cohort.

#### 3.1.2. Validation cohort

The validation cohort included 29 patients with pIBD and 10 non-IBD controls. Patient characteristics are shown in **Table 2**. The validation analyses were conducted on independent subjects except for the pIBD-JIA subgroup, for which the same 25 cases from the discovery cohort were reanalyzed, following re-extraction of RNA from tissue slides originating from the original FFPE mucosal biopsies.

**Table 2:** Patient characteristics of validation cohort.

|  |  | pIBD | pIBD-JIA | Non-IBD controls |
| --- | --- | --- | --- | --- |
| Patients | n | 29 | 25 | 10 |
| pCD/pUC |  | 13/16 | 10/15 |  |
| Age at pIBD diagnosis, years | median (IQR) | 13 (10-16) | 12 (11-15) | 7 (4-12) |
| Age at JIA diagnosis, years | median (IQR) |  | 14 (13-16) |  |
| Sex, females | n (%) | 11 (43) | 19 (76) | 5 (50) |

Abbreviations: pIBD, pediatric inflammatory bowel disease; IQR, interquartile range; JIA, juvenile idiopathic arthritis.

### 3.2. Discovery of miRNAs associated with JIA development in pIBD

MiRNA expression profiling with microarray analysis in the discovery cohort identified 532 probe sets across pIBD, pIBD-JIA, and non-IBD controls that were significantly differentially expressed between the groups (Figure 2). Expression values can be found in **Supplementary Table 2**. Of these, 98 probe sets were upregulated in patients with pIBD alone, relative with pIBD-JIA, whereas 434 were downregulated. Of these, 21 miRNAs had a fold change >3 while 123 miRNAs had a fold change >2 on a logarithmic scale (**Supplementary** Figure 1 & Figure 3).

**Figure 2.**
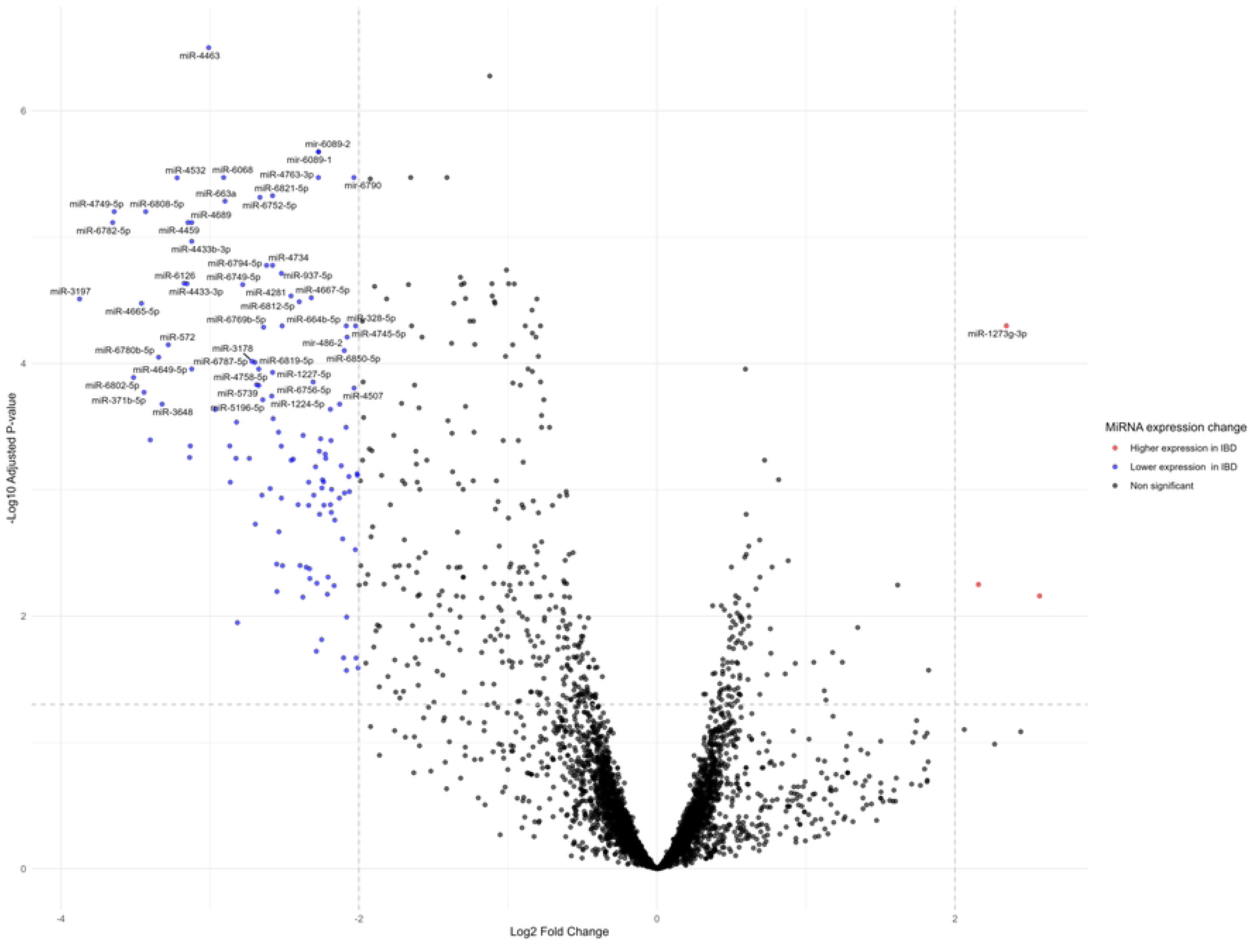
Volcano plot depicting differential miRNA expression in intestinal biopsies from patients with pediatric IBD (pIBD) compared to those with pIBD who later developed JIA (pIBD-JIA). Each point represents an individual miRNA, with the X-axis displaying the log₂ fold change and the Y-axis showing statistical significance as –log₁₀ of the FDR-adjusted p-value. Upregulated miRNAs in pIBD compared to pIBD-JIA are plotted on the right, while downregulated miRNAs are plotted on the left. Dashed lines indicate the significance threshold (adjusted p<0.05). pIBD, pediatric inflammatory bowel disease; pIBD-JIA, pediatric inflammatory bowel disease with later development of co-occurring juvenile idiopathic arthritis; non-IBD, non-inflammatory bowel disease; FFPE, formalin-fixed, paraffin-embedded; RT-qPCR, reverse transcription quantitative polymerase chain reaction.

**Figure 3.**
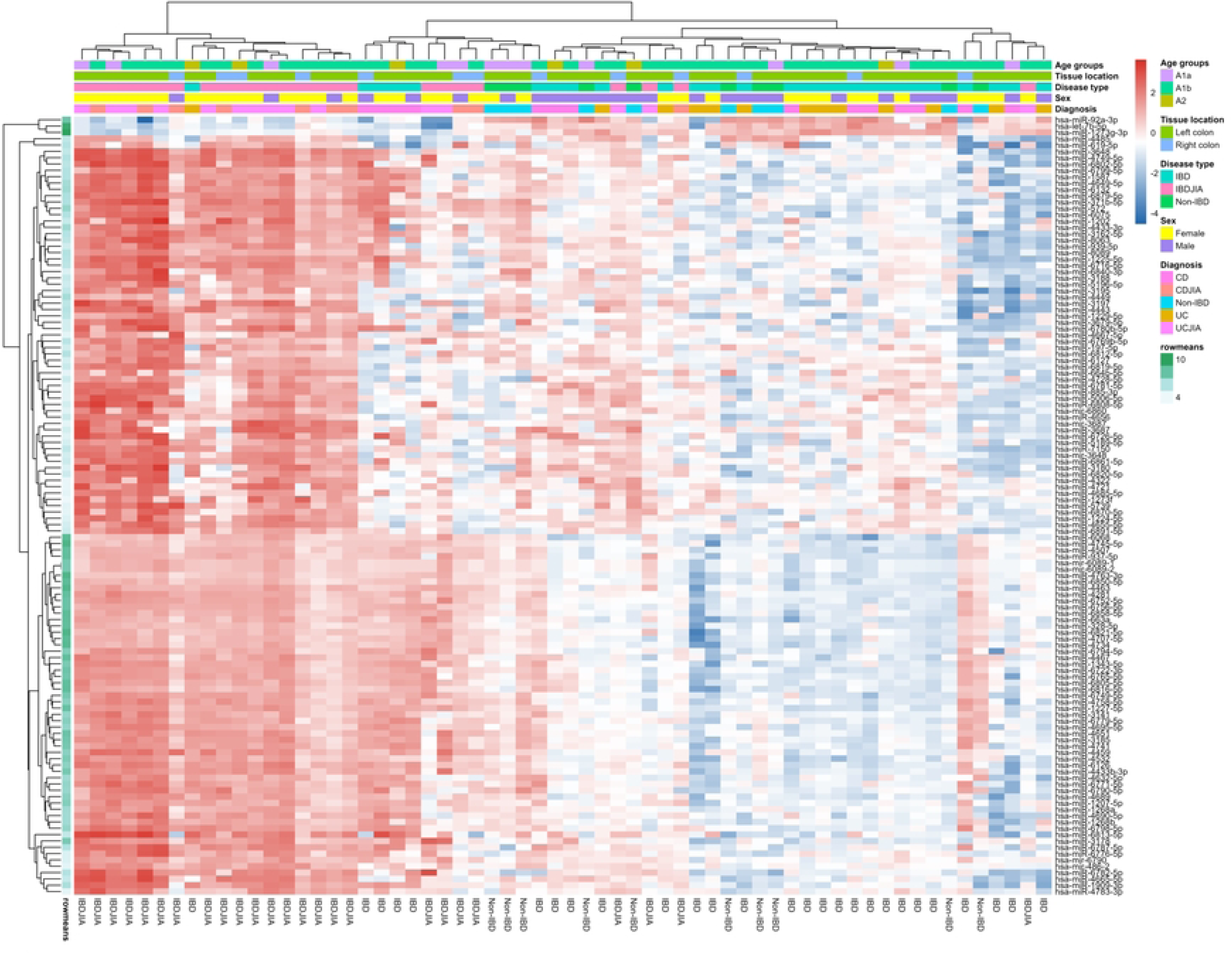
Heatmap of differentially expressed miRNAs between patients with pediatric IBD (pIBD) compared to those with pIBD who later developed JIA (pIBD-JIA). Heatmap displaying 123 significantly differentially expressed miRNAs (adjusted p < 0.05) between patients with pediatric inflammatory bowel disease (pIBD) and those who later developed juvenile idiopathic arthritis (pIBD-JIA), with an absolute log₂ fold change >2. Rows represent individual miRNAs, and columns represent patient samples. Expression values are scaled by row, with red indicating higher and blue indicating lower relative expression levels.

To further dissect miRNA expression patterns in relation to specific pIBD subtypes, we stratified the pIBD cohort into pCD and pUC groups and compared them to their respective pIBD-JIA counterparts. In pCD, 39 miRNAs were significantly upregulated compared to pCD-JIA (**Supplementary** Figure 2). Only miR-4443 had a log₂ fold change >2 (**Supplementary Table 3**). In contrast, pUC demonstrated a broader miRNA alteration, with 499 differentially expressed probe sets when comparing pUC and pUC-JIA (**Supplementary Table 4** & **Supplementary** Figure 3). Of these, 75 were upregulated and 424 were downregulated, with 12 miRNAs exhibiting a log₂ fold change >2.

No significant differences in miRNA expression were observed across sex (males vs. females), Paris age groups (A1a, A1b, or A2), histopathologic activity (RHI: remission, mild, or moderate-to-severe), clinical disease activity at diagnosis (abbrPCDAI/PUCAI: mild vs. moderate-to-severe), or sampling location (right vs. left colon) (**Supplementary Table 5).**

### 3.3. Clinical performance of miRNAs as biomarkers for JIA development in pIBD

A systematic visual review of expression plots was performed for all 123 miRNAs with a log₂ fold change of more than +/- 2 across pIBD and pIBD-JIA. Thirteen miRNAs—miR-3197, miR-6782-5p, miR-6808-5p, miR-619-5p, miR-3648, miR-572, miR-4433b-3p, miR-4463, miR-663a, miR-4281, miR-6891-5p, let-7b-5p, and miR-92a-3p—showed pronounced separation with minimal intergroup overlap in expression distributions (Figure 4). For these candidates, ROC analyses of pIBD vs. pIBD-JIA yielded AUCs above 0.7 for 11 miRNAs. Summary metrics are provided in **Supplementary Table 6**, with ROC curves shown in **Supplementary** Figure 4 (13 panels).

**Figure 4.**
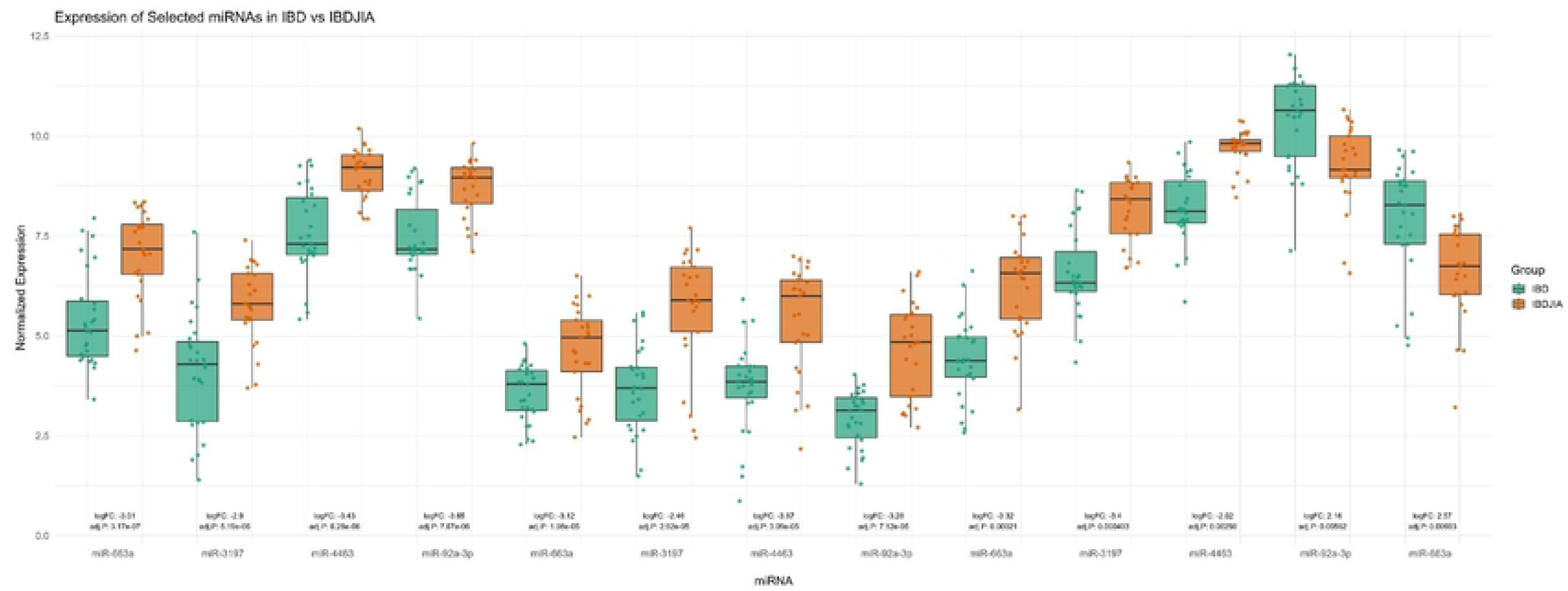
Box-plots of top candidate miRNAs distinguishing. patients with pediatric IBD (pIBD) from those with pIBD who later developed JIA (pIBD-JIA). Thirteen miRNAs (miR-3197, miR-6782-5p, miR-6808-5p, miR-619-5p, miR-3648, miR-572, miR-4433b-3p, miR-4463, miR-663a, miR-4281, miR-6891-5p, let-7b-5p, and miR-92a-3p) showing the most pronounced separation between pediatric IBD (pIBD) and pIBD with subsequent JIA (pIBD-JIA) in the discovery microarray.

### 3.4. Validation of miRNAs associated with JIA development in pIBD in an independent pIBD cohort

Four miRNAs—miR-663a, miR-3197, miR-4463, and miR-92a-3p—were selected for validation by RT-qPCR, as detailed in the methods section. ROC curves for the selected candidate miRNAs are shown in Figure 5. However, in the initial testing, the assays for mir-3197 and miR-4463 showed late Ct values for all samples and poor linearity, respectively, and were excluded from further analysis. Validation was conducted in the independent pIBD and non-IBD cohort; pIBD-JIA cases were identical to those in discovery (internal, non-independent validation). The UniSp6 spike in control added prior to RT was detected in all samples at comparable levels (Ct values between 20.6-21.8), indicating efficient cDNA synthesis for all samples. Following Ct value adjustment according to the IPC, expression of miR-663a and miR-92a-3p was determined relative to expression of reference miRNAs and the small nuclear RNAs U6 and SNORD48 which are commonly used for normalization in miRNA RT-qPCR-studies.^21^ When visually inspected, a remarkable overlap between patient groups were observed for both miR-663a and mir-92a-3p (Figure 6). Thus, neither of the candidates exhibiting differential expression could be reproduced with RT-qPCR in the validation cohort. To evaluate if this was related to assay technique or samples, analysis of RNA samples from the discovery cohort (six pUC and six pUC-JIA patients) was supplemented with qPCR which revealed differential expression between patient groups for both miR-663a (p<0.001) and miR-92a-3p (p<0.05), indicating that the methods are comparable but that the differentially expressed miRNAs were cohort-specific (**Supplementary** Figure 5).

**Figure 5.**
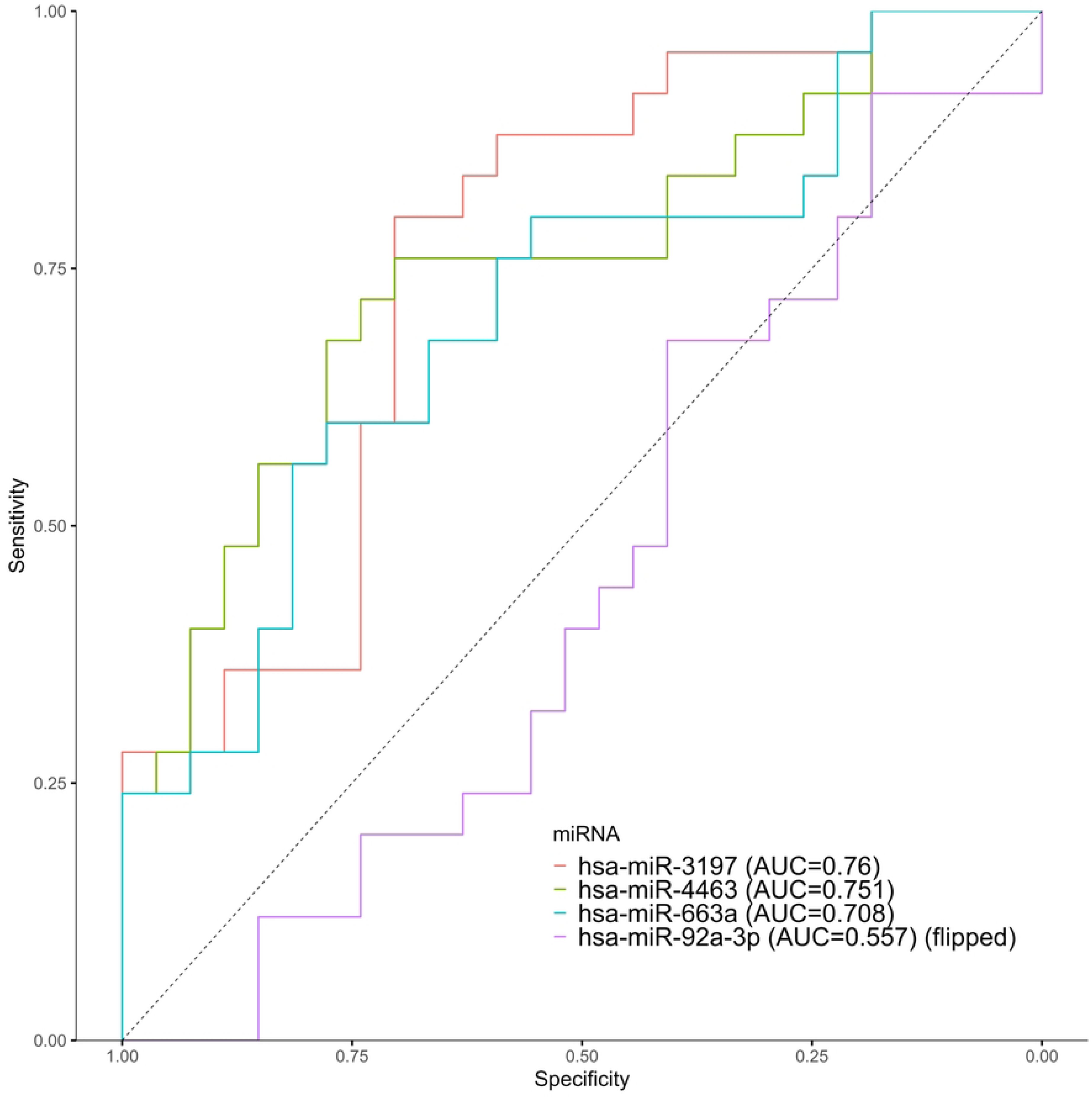
Receiver operating characteristic (ROC) curves for selected candidate miRNAs. ROC curves showing the discriminatory performance of the four selected miRNAs (miR-663a, miR-3197, miR-4463, and miR-92a-3p) in distinguishing patients with pediatric inflammatory bowel disease (pIBD) from those who later developed juvenile idiopathic arthritis (pIBD-JIA) in the discovery cohort. The area under the curve (AUC) quantifies classification performance for each miRNA.

**Figure 6.**
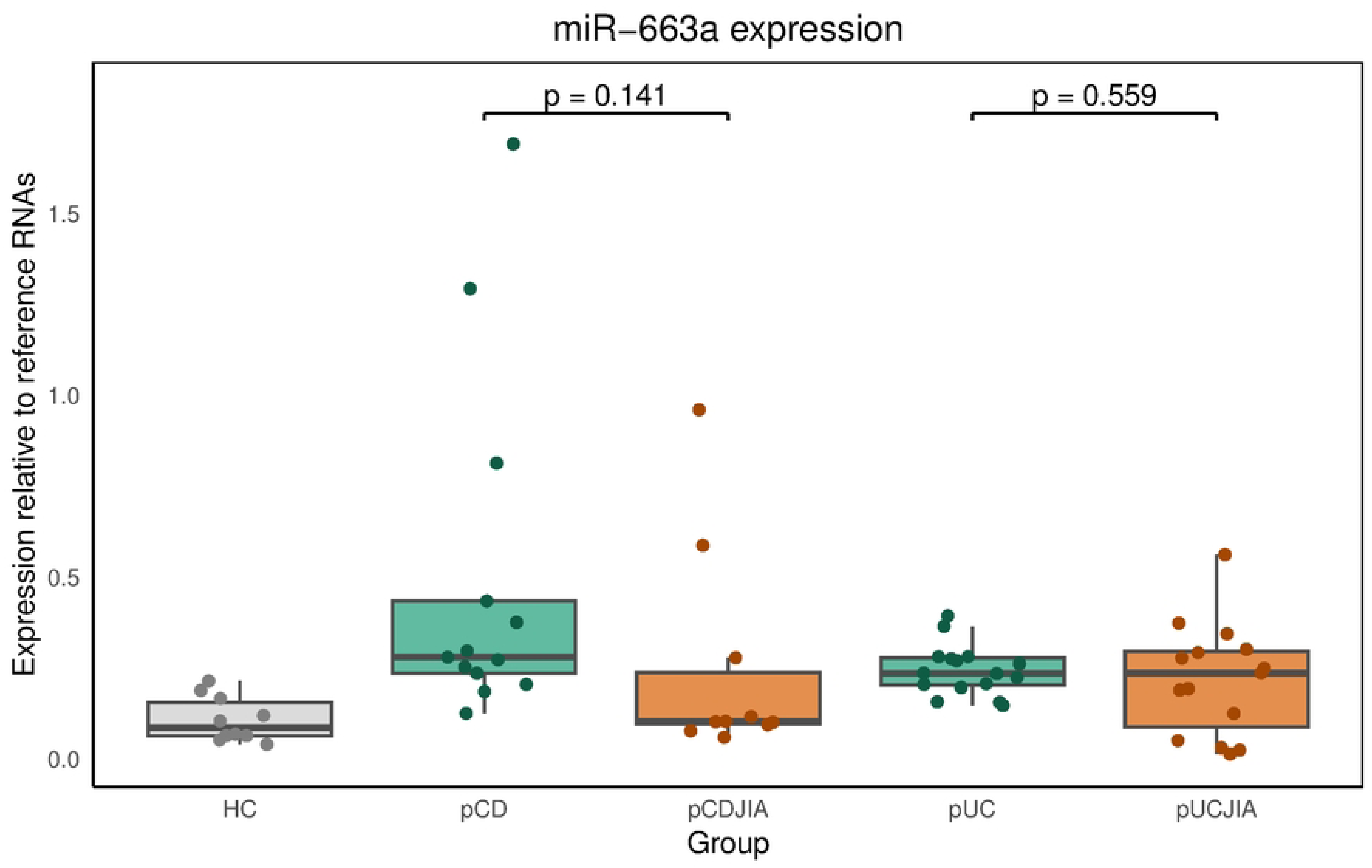
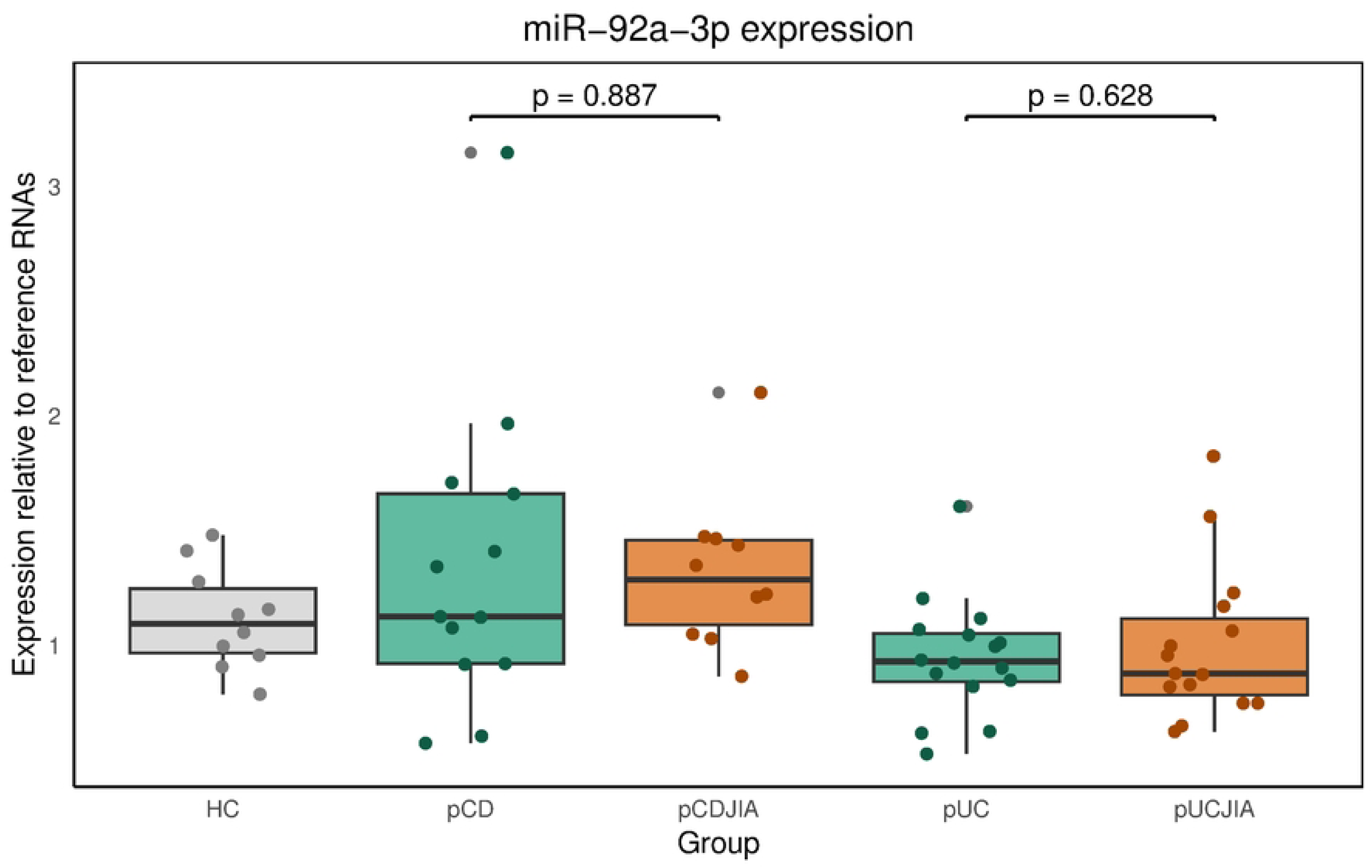
RT-qPCR validation of candidate microRNAs in the validation cohort. Relative expression of (A) miR-663a and (B) miR-92a-3p measured by RT-qPCR in pediatric IBD (pIBD) and pIBD with subsequent JIA (pIBD-JIA). Ct values were IPC-adjusted using UniSp6 and normalized (2^–ΔCt) to the average of miR-27a, miR-16-5p, miR-191-5p, U6, and SNORD48. Box-plots display median, interquartile range (IQR), and range. Visual overlap between groups was marked for both miRNAs, and no significant differences were detected in the validation cohort (Welch’s t-test, p>0.05).

## 4. DISCUSSION

### 4.1. Overview of main findings

In this study, diagnostic FFPE intestinal mucosal biopsies from the index endoscopy at pIBD diagnosis were subjected to microarray analysis to evaluate mucosal tissue miRNAs as potential prognostic biomarkers of subsequent JIA development prior to clinical onset. In the discovery study, microarray data yielded signatures that distinguished pIBD alone from pIBD cases who later developed JIA. From the microarray data, 13 candidate miRNAs were nominated and showed discrimination by visual inspection of expression and ROC analysis. Importantly, within the discovery cohort, relative expression of these candidates was replicated when quantified by RT-qPCR, with directionally consistent differences and similar relative separation between groups. However, when the selected miRNAs were tested in an independent pIBD/non-IBD cohort in the validation study, the differential expression was not reproduced. This lack of external replication underscores limited generalizability across cohorts and highlights the need for larger, independent populations before considering these miRNAs as robust biomarkers.

### 4.2. Cohort design and representativeness

The discovery and validation cohorts were designed to complement each other while reflecting the same underlying patient population. The discovery cohort included treatment-naïve patients with newly diagnosed pIBD, among whom a subset later developed JIA, enabling identification of mucosal miRNA patterns preceding joint disease onset. The validation cohort consisted of an independent group of treatment-naïve pIBD patients and non-IBD controls recruited at the same center and with comparable distributions of age, sex, and IBD subtype. Although the pIBD–JIA subgroup was reused due to the rarity of this condition, the close alignment between cohorts supports the representativeness of the validation cohort. Collecting such well-characterized, treatment-naïve biopsies from patients with pIBD, particularly those who subsequently develop JIA, is exceptionally challenging, emphasizing the uniqueness and value of this dataset.

### 4.3. Clinical context and relevance

This study addresses an unmet clinical need in pIBD. EIMs are common and often track with a more severe disease course; among them, JIA is particularly consequential, given its impact on pain, function, joint integrity, as well as psychosocial well-being. Prospective pediatric inception cohorts such as RISK^49^ (Crohn’s disease) and PROTECT^50^ (ulcerative colitis) have advanced risk stratification using clinical, serologic, microbial, and tissue molecular features, yet none provides a practical tool to predict JIA at the time of pIBD diagnosis. Although a small number of studies have profiled miRNA expression in pIBD or in JIA separately, to our knowledge no prior work has specifically interrogated miRNAs in patients with pIBD who subsequently develop concomitant JIA. **Supplementary Table 7** summarizes published studies profiling miRNAs in pIBD or JIA, using either intestinal mucosal biopsies or peripheral blood.

### 4.4. Candidate selection and biological interpretation

In the discovery study, a deliberately stringent selection threshold of log2 fold change > 2 was applied to ensure clear separation between diagnostic groups—stricter than the thresholds commonly used in microarray biomarker studies, including recent work in pIBD.^21,23^ Thirteen candidate miRNAs were nominated, and selection was further guided by prior literature linking several of these to inflammatory arthropathies. For example, miR-663a, miR-4281, and miR-6891 have been associated with immune and joint-related inflammatory pathways^51–53^, while let-7b and miR-92a-3p are known regulators of synovial and cartilage homeostasis in arthritis.^54,55^ The final validation panel included miR-663a and miR-92a-3p based on both biological relevance and assay feasibility.

Mechanistically, miR-663a shows context-dependent roles: in monocytes it suppresses AP-1 activity via JUNB/JUND (anti-inflammatory), whereas in rheumatoid-arthritis synovium it can activate canonical Wnt signaling through APC inhibition, promoting fibroblast proliferation.^56–58^ Conversely, miR-92a-3p (miR-17–92 cluster) supports cartilage-matrix maintenance via HDAC2 and WNT5A targeting and is typically reduced in arthritic tissue^59–61^, consistent with the lower levels observed in pIBD-JIA compared with pIBD alone in the discovery cohort. These multifunctional effects illustrate that miRNAs act through broad, interconnected networks, likely contributing to cohort-specific findings and the absence of replication across independent cohorts.^18^

### 4.5. Technical and analytical considerations

RT-qPCR reproduced the discovery-phase differences within a selected subset of patients from the discovery cohort. To maximize dynamic range, this subset was enriched for distributional extremes, and further analysis showed that the high fold changes were largely driven by nine pIBD patients.

These individuals differed in several clinical characteristics, and block age varied, underscoring how small groups can still disproportionately influence results. Despite cross-platform agreement, these findings did not validate externally, emphasizing the importance of adequately powered, independent replication.

Joint manifestations remain the most frequent EIM in pIBD but still affect a minority of patients overall. In Danish population-based data, approximately 4% had an EIM at diagnosis and an additional 14% during follow-up, with joint involvement among the most common.^5^ Similar frequencies have been reported internationally.^62,63^ These epidemiological and practical constraints, together with limited availability of archived diagnostic tissue, justified re-use of the rare pIBD-JIA cases for assay validation.

Archived FFPE intestinal biopsies were therefore the most feasible material for retrospective studies. Although RNA yield is lower than from fresh-frozen tissue, miRNAs are known to be resilient to fixation and long-term storage, and their profiles correlate well between FFPE and frozen samples.^65–67^ In our cohort, no decline in global mean miRNA expression was observed with block age (**Supplementary** Figure 6), supporting the analytical suitability of FFPE for miRNA-based studies in this rare phenotype.

### 4.6. Strengths and limitations

A key strength of this study is the relatively large discovery cohort for this rare phenotype: 62 diagnostic intestinal biopsies profiled on the Affymetrix GeneChip miRNA 4.0 platform. Most prior microarray studies in pIBD, JIA, or related autoimmune conditions included fewer samples (often <30) or narrower assay content, limiting discovery power. Further strengths include clinically verified, treatment-naïve pIBD-JIA cases and use of two analytic platforms (microarray followed by RT-qPCR), which reduces platform-specific bias and strengthens confidence in any recurring signals.

Limitations include a small validation cohort and non-independent pIBD-JIA cases at validation, constraining external generalizability. Although miRNAs are relatively stable in fixed tissue, reliance on archived FFPE (rather than fresh-frozen) can introduce pre-analytical variability. The low prevalence of pIBD-JIA also means a few patients can disproportionately influence group differences, as seen in our enriched-subset analysis. Finally, unmeasured confounding (e.g., subtle pre-analytical handling differences) may contribute to inter-cohort discrepancies; we mitigated this by interleaving samples across runs and applying ComBat batch correction, yet residual effects cannot be fully excluded.

### 4.7. Conclusions and future directions

This study evaluated whether mucosal miRNAs measured at the time of pIBD diagnosis can predict subsequent development of JIA. Using diagnostic FFPE intestinal biopsies, we performed microarray profiling in a discovery cohort (n=62; pIBD, pIBD–JIA, and non-IBD controls) and then selected candidates for verification by RT-qPCR, followed by validation in an independent pIBD/non-IBD cohort (with pIBD–JIA cases reused due to rarity). However, these signals did not replicate in the validation cohort, indicating that cross-cohort generalizability is unproven and that the observed markers should be considered exploratory.

Taken together, our findings suggest that mucosal miRNAs capture biology relevant to joint involvement in pIBD but, as currently defined, lack the robustness required for clinical translation. The discrepancy between discovery and validation likely reflects small sample sizes in a rare phenotype, cohort-specific heterogeneity, and pre-analytical/analytical variability inherent to archived tissue. Future work should prioritize larger, fully independent multicenter cohorts, standardized pre-analytical handling and shared analysis pipelines, longitudinal sampling from diagnosis through follow-up, and replication across platforms and specimen types (tissue and blood). Only with such rigor will it be possible to determine whether mucosal miRNA profiles at pIBD onset can reliably stratify risk for later JIA and support clinically actionable prognostic tools.

## Data Availability

Microarray expression data have been deposited in ArrayExpress at the European Bioinformatics Institute (EMBL-EBI) under the accession number E-MTAB-15807.

https://www.ebi.ac.uk/

## ACKNOWLEDGMENTS

We thank all children and adolescents who contributed biopsies to this project. We also acknowledge the dedication of the endoscopy unit staff at the Gastrounit, Copenhagen University Hospital, Amager and Hvidovre, Denmark, for their support in biopsy collection for the ongoing Copenhagen IBD Inception Cohort Study. Additionally, we appreciate the assistance of laboratory technicians in RNA extraction and microarray analyses, as well as molecular biologists in RT-qPCR validation. Finally, we acknowledge the contribution of pathologists from Copenhagen University Hospital, Herlev and Gentofte, Denmark, in selecting FFPE blocks through histopathological review.

